# Changing Faces of Authorship: A Study of Gender, Race, Regional Disparities in Gastroenterology/Hepatology-related RCTs. A Two Decade Analysis (2000-2022)

**DOI:** 10.1101/2023.08.28.23294747

**Authors:** Roopa Kumari, FNU Sadarat, Sindhu Luhana, Om Parkash, Subhash Chander

## Abstract

**Objective:** To investigate gender, racial, ethnic, and regional disparities in first and senior authorship positions in gastroenterology/hepatology-related randomised controlled trials (RCT).

**Design:** Retrospective bibliometric analysis of PubMed-indexed RCTs published between January 2000 to December 2022 in leading journals with an impact factor of at least five.

**Results:** 943 RCTs met our inclusion criteria, providing a participant pool of 301 female (15.96%) and 1,585 male (84.04%) authors from 37 countries (70% high-income countries). Despite a significant increase in the proportion of female authors in first and senior authorship positions between 2000 and 2022 (*p*<0.001), females were grossly underrepresented in both authorship positions, with a male-to-female ratio of 4.45 and 6.37, respectively. The male-to-female ratio was highest among Asian authors (7.79) than among White (4.22), Hispanic (1.44), and Black (1) authors in the first authorship position. In contrast, the male-to-female ratio was similar for Asian (6.2) and White (6.67) authors in the senior authorship position, with a low underlying frequency of Hispanic and Black female authors. There were statistically significant differences in gender distribution for first authorship at the country level (*p*=0.0018). Binary logistic regression analysis showed significant positive effects of the senior author being a female (β=1.124, p<0.001) and the senior author having PhD qualification (β=0.753, p=0.021) on female first authorship.

**Conclusion:** Despite significant improvements in gender, racial and ethnic representation in first and senior authorship of gastroenterology/hepatology-related RCTs published in high-impact journals, progress toward parity remains slow. Targeted interventions to improve author diversity are warranted.

## INTRODUCTION

Females and racial/ethnic minorities are underrepresented as authors in medical research publications, particularly in first and senior authorship positions commonly associated with increased recognition and career advancement [1, 2, 3]. In addition, several studies have shown that the gender and racial/ethnic disparities in research output may be more pervasive in surgical and procedural fields such as gastroenterology and hepatology [4, 5, 6, 7, 8]. For instance, an analysis of 114 academic gastroenterology programs in the United States showed that male faculty had longer careers with better publication track records and higher h-indices, which correlated with higher academic rank [9].

Similarly, gender disparities have been noted in the proportion of females in senior faculty positions. For example, in 2014, less than a quarter of faculty in academic gastroenterology programs in the United States were females, of which only 29% held senior faculty positions compared to 50% of males [9]. John *et al.* [10] reported similar trends in the proportion and productivity of female gastroenterologists in 2017. Additionally, gender disparities have been reported in chief editorship and editorial board membership of gastroenterology and hepatology journals [6], as program director, associate program director, division chief or chairs of gastroenterology fellowship programs [11, 12, 13], and as speakers or chairs of major gastroenterology conferences [14].

The underrepresentation of females in gastroenterology and hepatology has been attributed to personal choice [15], lack of female role models or mentors [16], gender bias at the workplace [16, 17], and most commonly to the challenges in balancing work-life commitments such as the overlap of fellowship years with planned parenthood, long work week, and frequent night calls [4, 16, 18, 19]. For instance, Singh *et al.* [20] demonstrated that after ten years of clinical practice, female gastroenterologists have fewer children, are paid 22% less, and hold fewer leadership positions than their male counterparts. Furthermore, a more recent report showed that female gastroenterologists were paid 10-20% less than their male counterparts in 2018-2019, highlighting the increasing pay gap with higher academic ranks [21]. Together these studies suggest that the progress towards gender parity in gastroenterology and hepatology, if any, has been slow in the past decade.

Racial and ethnic disparities in the authorship of gastroenterology and hepatology journal articles have been relatively less studied than gender disparities. However, data from Accreditation Council for Graduate Medical Education (ACGME) showed that of the 1,911 active gastroenterology residents during 2021-2022, 37.6% self-identified as White and 40.2% as Asian, while only 7.6% and 5.5% identified as Hispanic or Black [22]. In addition, a recent survey of gastroenterology and hepatology medical professionals in the United States by Rahal *et al.* [23] identified the underrepresentation of racial and ethnic minority groups in the medical education/training, practising gastroenterology and hepatology professionals in the workplace, and professional leadership positions as barriers to increasing racial and ethnic diversity. Given the underrepresentation of racial and ethnic minority groups in the gastroenterology and hepatology workforce, it is highly likely that racial and ethnic disparities in authorship are as prevalent, if not worse, as gender disparities.

Furthermore, earlier studies have noted that the proportion of female authors in medical research varies by region, with a higher proportion of females in publications coming out of institutions in Oceania, Europe, and North America than those from Asia, South America, and Africa [24, 25]. Consistent with these trends, others have shown gender, racial or ethnic underrepresentation and fewer journal submissions from low-and middle-income countries [26, 27, 28]. Therefore, in this bibliometric study, we investigated the gender, racial, ethnic, and regional disparities in first and senior authorship positions among gastroenterology and hepatology articles published in thirteen leading speciality and general medical journals. There is also good evidence that the proportion of females may vary by study design [24, 25]. For instance, a lower proportion of female authors were reported in experimental studies (34.2%), systematic reviews (41.8%), and cross-sectional, cohort or case-control studies (45.5) than in qualitative research (74%) [24]. Given the significance of randomised controlled trials (RCTs) in guiding clinical practice and decision-making, we limited our bibliometric analysis to gastroenterology/hepatology-related RCTs published between 2000 and 2022.

## METHODS

### Study Design

A comprehensive retrospective analysis of PubMed-indexed RCTs in gastroenterology and hepatology journals was conducted from January 2000 to December 2022. The following Medical Subject Headings (MeSH) search terms were used in the PubMed search: “gastroenterology,” “hepatology,” “randomised controlled trials,” “authorship,” “gender,” and “race and health.” Search strings were created to capture all relevant articles, combining terms with Boolean operators as follows: ((gastroenterology OR hepatology) AND (randomised controlled trials) AND (authorship) AND (gender OR race and health)). Filters for publication dates (01-01-2000 to 12-31-2022) and language (English) were also applied.

For this study, journals with an impact factor of 5 or above were classified as high-impact journals. Accordingly, the following high-impact journals were screened for gastroenterology/hepatology-related RCTs: BMJ, Clinical Gastroenterology and Hepatology, Gastroenterology, Gut, Gut Microbes, The Journal of the American Medical Association (JAMA), Journal of Gastroenterology, Journal of Hepatology, Lancet, Microbiome, Nature, The New England Journal of Medicine (NEJM), and The American Journal of Gastroenterology.

### Inclusion and Exclusion Criteria

All RCTs published in the thirteen selected journals in the English language focusing on gastroenterology and hepatology-related topics, including but not limited to inflammatory bowel diseases, functional gastrointestinal disorders, liver diseases, gastrointestinal cancers, hepatitis, and gastrointestinal endoscopy, were considered for inclusion.

Articles published in a non-English language, article full-text unavailable through the journal homepage or an online repository, non-RCT studies, including case reports, case series, retrospective studies, cross-sectional studies, cohort studies, review articles, commentaries, editorials, letters to the editor, and any articles published in journals other than the high-impact gastroenterology and hepatology journals were excluded from this study. Additionally, studies with unclear or missing information on first and/or senior authorship and those for which gender or race/ethnicity cannot be reasonably determined even after attempting to contact the authors were excluded. Finally, studies on topics unrelated to gastroenterology or hepatology, such as general internal medicine, surgery, or radiology, were excluded.

### Data Extraction

The data extraction process was carried out in three steps. First, a list of PubMed-indexed RCTs published during the study period was generated. Second, the RCTs were filtered based on the journal’s impact factor. Finally, authorship-related data were extracted from articles meeting our inclusion criteria using a standardised form.

For gender and racial/ethnic data of first and senior authors, we used a combination of name-based algorithms such as Genderize and Ethnea. Discrepancies or ambiguities were resolved using manual online searches of the author’s institutional websites, biographies, and social media profiles. In cases where the information was not readily available or was ambiguous, we contacted the authors via email for self-identification.

Race/ethnicity was categorised into five groups: White, Asian, Black, Hispanic, and others. The “other” category included individuals who did not fit the categorisation or identified as multiracial or multi-ethnic. In addition to gender and race/ethnicity, data related to the educational attainment of the authors and the country of the author’s affiliated institutions were also extracted. Countries were categorised by region and income level according to the 2023 World Bank classification [29].

### Statistical Analysis

Descriptive statistics, including frequency, percentage and male-to-female ratio, were used to summarise the distribution of gender and race/ethnicity across first and senior authorship positions. We also performed chi-squared tests to examine the association between journals and gender, gender and educational attainment, journals and race/ethnicity, and country of affiliation and gender or race/ethnicity of the first and senior authors.

To analyse the association between gender and race/ethnicity with first and senior authorship, we performed a logistic regression analysis, adjusting for potential confounders such as year of publication, country of origin, educational attainment, and journal name. The results are presented as odds ratios (ORs) with 95% confidence intervals (CIs). Nagelkerke R Square and Cox and Snell R Square were calculated to determine the first and senior authorship variability. All analyses were performed using R software 4.2.0 [30], with a two-sided *p*-value of <0.05 considered statistically significant.

## RESULTS

### Descriptive Statistics of the Study Population

943 RCTs met our inclusion criteria, providing a participant pool of 301 female (15.96%) and 1,585 male (84.04%) authors for this study (**Table 1**). Nearly 80% of the study population was White, and 89.7% came from high-income countries, especially Europe & Central Asia (41.36%) and North America (37.12%). The Gastroenterology journal contributed the highest portion of RCTs (16.65%), followed by Am J Gastroenterol. (16.54%), Lancet (13.57%), Gut (11.24%), BMJ (11.03%), Clin Gastroenterol Hepatol. (9.01%), NEJM (7%), JAMA (5.41%), J Hepatol. (2.65%), J Gastroenterol. (2.55%), Nature (2.55%), Gut microbes (1.48%), and Microbiome (0.32%).

**Table 1:** Frequency and proportion of female authors in first and senior authorship positions with male-to-female ratio (M:F) by race, ethnicity, educational attainment, and World Bank income and region groups.

| Variables | First Authorship |  |  | Senior Authorship |  |  |
| --- | --- | --- | --- | --- | --- | --- |
|  | Female<br>n (%) | M:F | *p* | Female<br>n (%) | M:F | *p* |
| *Overall* | 173 (18.35%) | 4.45 | - | 128<br>(13.57%) | 6.37 |  |
| **Race and Ethnicity** |  |  |  |  |  |  |
| *White* | 144 (19.15%) | 4.22 | 0.002 | 105<br>(13.89%) | 6.2 | 0.820 |
| *Asian* | 19 (11.38%) | 7.79 |  | 21 (13.04%) | 6.67 |  |
| *Hispanic* | 9 (40.91%) | 1.44 |  | 2 (8%) | 11.5 |  |
| *Black* | 1 (50%) | 1 |  | 0 (0%) | (*Note 1*) |  |
| **Education Attainment** |  |  |  |  |  |  |
| *MD* | 120 (16.51%) | 5.06 | 0.005 | 98 (13.39%) | 6.47 | 0.245 |
| *Master's* | 1 (50%) | 1 |  | 0 (0%) | (*Note 2*) |  |
| *Master's, PhD* | 1 (100%) | 0 |  | 0 (0%) | (*Note 3*) |  |
| *MD, Master's* | 12 (25%) | 3 |  | 8 (27.59%) | 2.63 |  |
| *MD, Master's, PhD* | 0 (0%) | (*Note 4*) |  | 0 (0%) | (*Note 4*) |  |
| *MD, PhD* | 22 (19.13%) | 4.23 |  | 12 (9.76%) | 9.25 |  |
| *PhD* | 17 (35.42%) | 1.82 |  | 10 (17.54%) | 4.7 |  |
| **World Bank Income Group** |  |  |  |  |  |  |
| *High* | 162 (19.13%) | 4.23 | 0.123 | 112<br>(13.25%) | 6.54 | 0.131 |
| *Upper-middle* | 10 (13.33%) | 6.5 |  | 15 (19.74%) | 4.07 |  |
| *Lower-middle* | 1 (4.76%) | 20 |  | 1 (4.55%) | 21 |  |
| **World Bank Region** |  |  |  |  |  |  |
| *Latin America & Caribbean* | 0 (0%) | (*Note 5*) | 0.091 | 0 (0%) | (*Note 5*) | 0.195 |
| *East Asia & Pacific* | 25 (14.71%) | 5.8 |  | 27 (16.17%) | 5.19 |  |
| *Europe & Central Asia* | 85 (22.14%) | 3.52 |  | 42 (10.61%) | 8.43 |  |
| *South Asia* | 1 (5.26%) | 18 |  | 1 (5%) | 19 |  |
| *North America* | 62 (17.46%) | 4.73 |  | 57 (16.52%) | 5.05 |  |
| *Sub-Saharan Africa* | 0 (0%) | (*Note 5*) |  | 0 (0%) | (*Note 5*) |  |
| *Middle East & North Africa* | 0 (0%) | (*Note 5*) |  | 1 (10%) | 9 |  |

Notes:
(Note 1): Only one male and no female Black authors were in the senior authorship position.
(Note 2): No senior author reported only a Master's degree.
(Note 3): There were only two senior authors, both males, who reported a "Master's, PhD" degree.
(Note 4): There were only two first authors (both males) and no senior authors for either gender who reported an "MD, Master's, PhD" degree.
(Note 5): There were three authors from Latin America & Caribbean, one from Sub-Saharan Africa, and eleven from the Middle East & North Africa regions in the first authorship position, all males. Similarly, there were four male authors from Latin America & Caribbean and one from Sub-Saharan Africa in senior authorship positions with no female authors.

### Gender and Racial Disparities in First and Senior Authorship Positions

There was a significant increase in the proportion of female authors in first and senior authorship positions from 5% and 2.5% in 2000 to 20.48% in 2022 (*p*<0.001; **Table 2**). As a result, the male-to-female ratio decreased from 19 to 3.88 for the first authorship position and 39 to 3.88 for the senior authorship position during the study period (**Figure 1A**). However, despite the improving trend, females were grossly underrepresented in both first (18.35%) and senior (13.57%) authorship positions during the 22 years, with a male-to-female ratio of 4.45 and 6.37, respectively (**Table 1**).

**Table 2:**
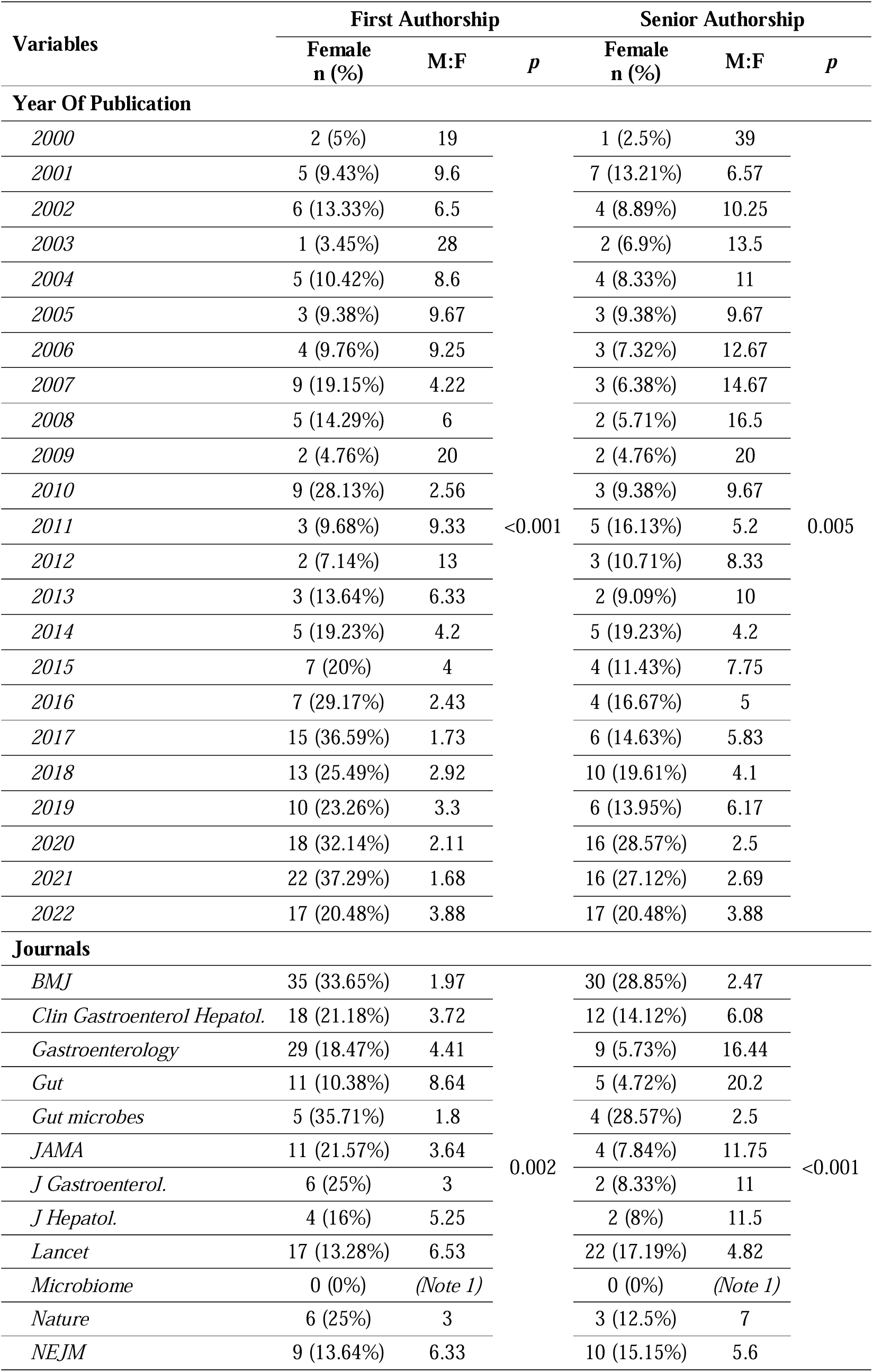
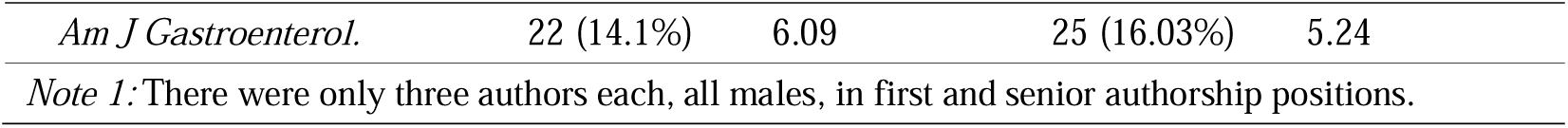
Frequency and proportion of female authors in first and senior authorship positions with male-to-female ratio (M:F) by the year and journal of publication.

| Variables | First Authorship |  |  | Senior Authorship |  |  |
| --- | --- | --- | --- | --- | --- | --- |
|  | Female<br>n (%) | M:F | *p* | Female<br>n (%) | M:F | *p* |
| Year Of Publication |  |  |  |  |  |  |
| 2000 | 2 (5%) | 19 | <0.001 | 1 (2.5%) | 39 | 0.005 |
| 2001 | 5 (9.43%) | 9.6 |  | 7 (13.21%) | 6.57 |  |
| 2002 | 6 (13.33%) | 6.5 |  | 4 (8.89%) | 10.25 |  |
| 2003 | 1 (3.45%) | 28 |  | 2 (6.9%) | 13.5 |  |
| 2004 | 5 (10.42%) | 8.6 |  | 4 (8.33%) | 11 |  |
| 2005 | 3 (9.38%) | 9.67 |  | 3 (9.38%) | 9.67 |  |
| 2006 | 4 (9.76%) | 9.25 |  | 3 (7.32%) | 12.67 |  |
| 2007 | 9 (19.15%) | 4.22 |  | 3 (6.38%) | 14.67 |  |
| 2008 | 5 (14.29%) | 6 |  | 2 (5.71%) | 16.5 |  |
| 2009 | 2 (4.76%) | 20 |  | 2 (4.76%) | 20 |  |
| 2010 | 9 (28.13%) | 2.56 |  | 3 (9.38%) | 9.67 |  |
| 2011 | 3 (9.68%) | 9.33 |  | 5 (16.13%) | 5.2 |  |
| 2012 | 2 (7.14%) | 13 |  | 3 (10.71%) | 8.33 |  |
| 2013 | 3 (13.64%) | 6.33 |  | 2 (9.09%) | 10 |  |
| 2014 | 5 (19.23%) | 4.2 |  | 5 (19.23%) | 4.2 |  |
| 2015 | 7 (20%) | 4 |  | 4 (11.43%) | 7.75 |  |
| 2016 | 7 (29.17%) | 2.43 |  | 4 (16.67%) | 5 |  |
| 2017 | 15 (36.59%) | 1.73 |  | 6 (14.63%) | 5.83 |  |
| 2018 | 13 (25.49%) | 2.92 |  | 10 (19.61%) | 4.1 |  |
| 2019 | 10 (23.26%) | 3.3 |  | 6 (13.95%) | 6.17 |  |
| 2020 | 18 (32.14%) | 2.11 |  | 16 (28.57%) | 2.5 |  |
| 2021 | 22 (37.29%) | 1.68 |  | 16 (27.12%) | 2.69 |  |
| 2022 | 17 (20.48%) | 3.88 |  | 17 (20.48%) | 3.88 |  |
| Journals |  |  |  |  |  |  |
| BMJ | 35 (33.65%) | 1.97 | 0.002 | 30 (28.85%) | 2.47 | <0.001 |
| Clin Gastroenterol Hepatol. | 18 (21.18%) | 3.72 |  | 12 (14.12%) | 6.08 |  |
| Gastroenterology | 29 (18.47%) | 4.41 |  | 9 (5.73%) | 16.44 |  |
| Gut | 11 (10.38%) | 8.64 |  | 5 (4.72%) | 20.2 |  |
| Gut microbes | 5 (35.71%) | 1.8 |  | 4 (28.57%) | 2.5 |  |
| JAMA | 11 (21.57%) | 3.64 |  | 4 (7.84%) | 11.75 |  |
| J Gastroenterol. | 6 (25%) | 3 |  | 2 (8.33%) | 11 |  |
| J Hepatol. | 4 (16%) | 5.25 |  | 2 (8%) | 11.5 |  |
| Lancet | 17 (13.28%) | 6.53 |  | 22 (17.19%) | 4.82 |  |
| Microbiome | 0 (0%) | (Note 1) |  | 0 (0%) | (Note 1) |  |
| Nature | 6 (25%) | 3 |  | 3 (12.5%) | 7 |  |
| NEJM | 9 (13.64%) | 6.33 |  | 10 (15.15%) | 5.6 |  |

|  |  |  |  |  |
| --- | --- | --- | --- | --- |
| *Am J Gastroenterol.* | 22 (14.1%) | 6.09 | 25 (16.03%) | 5.24 |
| *Note 1:* There were only three authors each, all males, in first and senior authorship positions. |  |  |  |  |

**Figure 1:**
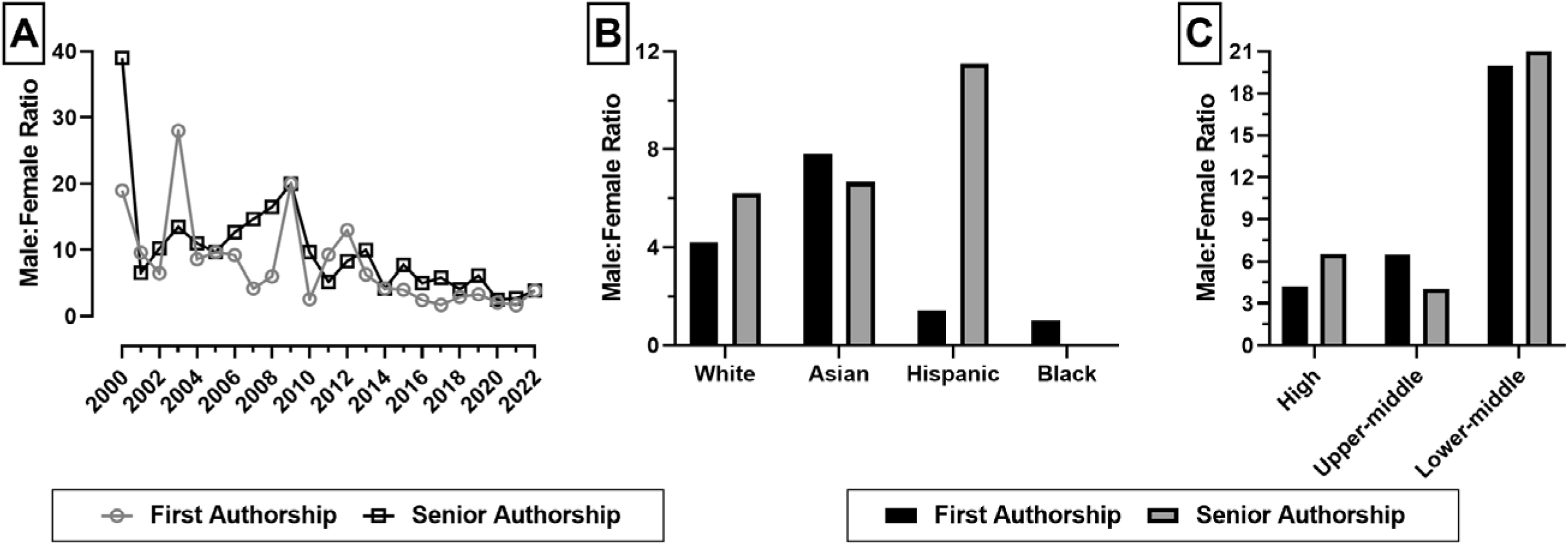
Male-to-female ratio in first and senior authorship positions by (A) the year of publication, (B) race or ethnicity of the authors, and the World Rank region the author’s affiliated institution

We also noted racial disparities within the pool of female authors, which aligned with the overall study population; 83.24% and 82.03% of all female authors in the first and senior authorship positions identified as White, compared to 79.96% of all authors in the overall study population (data not shown). However, differences were noted in the proportion of female authors within each race/ethnic group, with statistical significance only for the first authorship position (*p*=0.002, Table 1). For example, the male-to-female ratio was highest among Asian authors (7.79) than among White (4.22), Hispanic (1.44), and Black (1) authors in the first authorship position (**Figure 1B**, **Table 1**). In contrast, the male-to-female ratio was similar for Asian (6.2) and White (6.67) authors in the senior authorship position, with only a few Hispanics (n=8; male-to-female ratio=11.5) and no Black female authors (**Figure 1B**, **Table 1**). A significant relationship was found between the year of publications and the race of the first author [χ2(66, N = 943) = 139.55, *p* = 0.028] (data not shown).

Regarding educational attainment, only 35% of the first authors with a PhD, 25% with MD+Master’s, 16.51% with MD, and 19.13% with MD+PhD degrees were females. Similarly, female authors comprised 27.59% of senior authors with MD+Master’s, 17.54% with PhD, 13.39% with MD, and 9.64% with MD+PhD degrees (**Table 1**). There was a significant positive association between the gender of the first and senior authors [χ2(1, N = 943) = 42.43, *p* < 0.001] and the education of the first and senior authors [χ2(24, N = 943) = 47.95, *p* < 0.001] (data not shown).

Among journals, Gut microbes (1.8 & 2.5) and BMJ (1.97 & 2.47) had the lowest male-to-female ratio, while Gut (8.64 & 20.2) had the highest in both first and senior author positions (**Table 2**). In addition, there was a statistically significant association between the gender of the first (*p*=0.002) or senior (*p*<0.001) author and the journal (**Table 2**).

### Region Disparities in First and Senior Authorship Positions

The majority of the first authors in our study population were affiliated with institutions in high-income countries (n=26 countries), followed by upper-middle-income (n=6) and lower-middle countries (n=5). Although the male-to-female ratio for the first authorship position decreased numerically with income, the difference was insignificant (**Figure 1C**, **Table 1**; *p*=0.123). Among World Bank regions, Europe & Central Asia (3.52) had the lowest male-to-female ratio, followed by North America (4.73), East Asia & Pacific (5.8), and South Asia (18) (**Table 1**; *p*=0.091).

In contrast, the male-to-female ratio was the lowest in upper-middle-income (4.07) compared to high-income (6.54) and upper-middle-income (21) countries in the senior authorship position, although the difference did not reach statistical significance (**Figure 1C**, **Table 1**; *p*=0.131). Among World Bank regions, North America (5.05) and East Asia & Pacific (5.19) had numerically lower male-to-female ratios than Europe & Central Asia (8.43) and South Asia (19) (**Table 1**; *p*=0.195).

However, statistically significant differences in gender distribution in the first but not the second authorship position were noted by country (*p*=0.0018; **Table 3**), with the largest number of female authors in the first authorship position affiliated with institutions in the United States (n=51, 29.48%; **Table 1**).

**Table 3:**
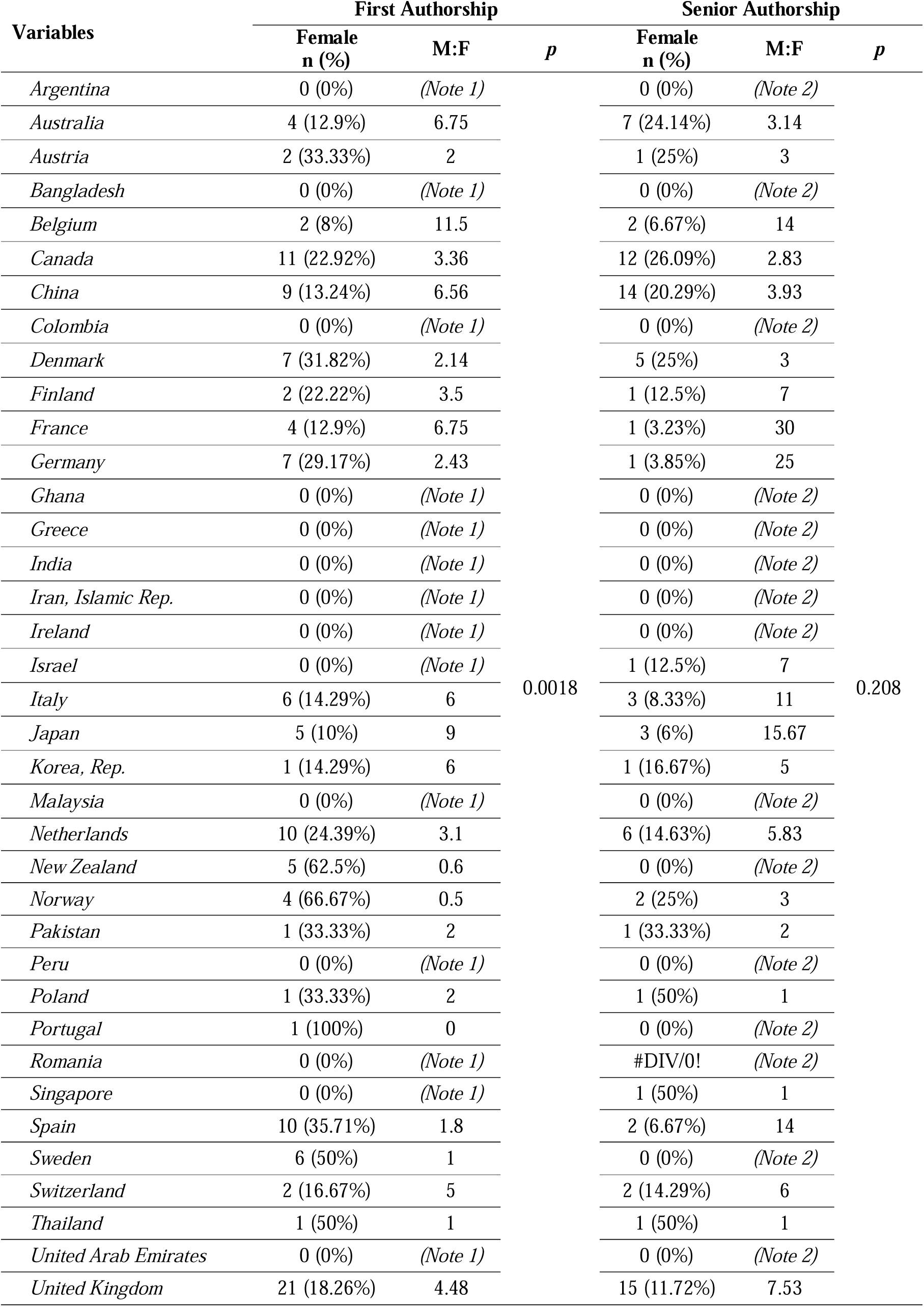
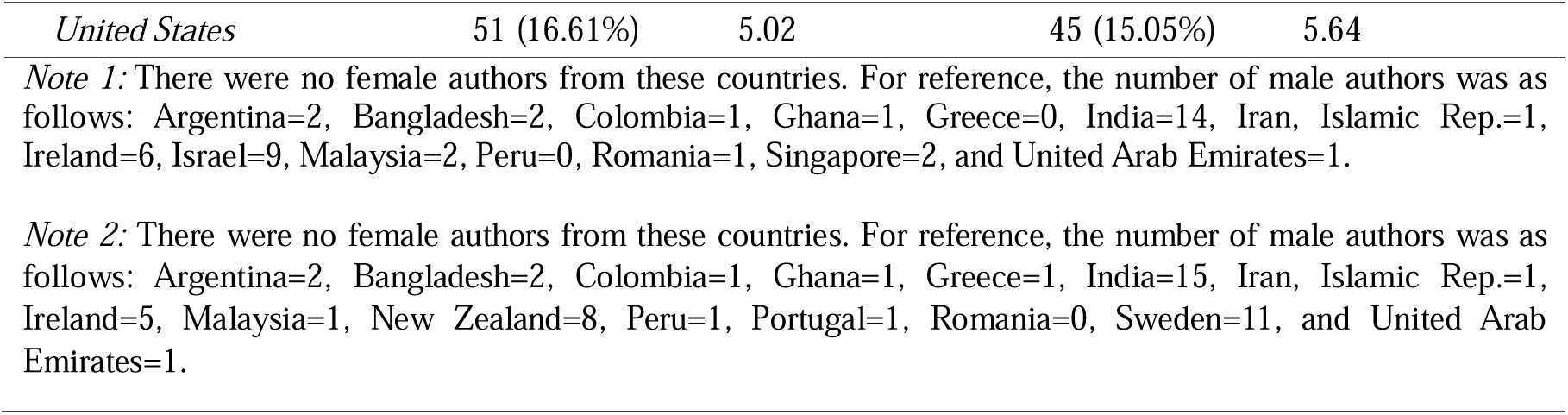
Frequency and proportion of female authors in first and senior authorship positions with male-to-female ratio (M:F) by the country of affiliated institution.

| Variables | First Authorship |  | *p* | Senior Authorship |  | *p* |
| --- | --- | --- | --- | --- | --- | --- |
|  | Female<br>n (%) | M:F |  | Female<br>n (%) | M:F |  |
| *Argentina* | 0 (0%) | (Note 1) | 0.0018 | 0 (0%) | (Note 2) | 0.208 |
| *Australia* | 4 (12.9%) | 6.75 |  | 7 (24.14%) | 3.14 |  |
| *Austria* | 2 (33.33%) | 2 |  | 1 (25%) | 3 |  |
| *Bangladesh* | 0 (0%) | (Note 1) |  | 0 (0%) | (Note 2) |  |
| *Belgium* | 2 (8%) | 11.5 |  | 2 (6.67%) | 14 |  |
| *Canada* | 11 (22.92%) | 3.36 |  | 12 (26.09%) | 2.83 |  |
| *China* | 9 (13.24%) | 6.56 |  | 14 (20.29%) | 3.93 |  |
| *Colombia* | 0 (0%) | (Note 1) |  | 0 (0%) | (Note 2) |  |
| *Denmark* | 7 (31.82%) | 2.14 |  | 5 (25%) | 3 |  |
| *Finland* | 2 (22.22%) | 3.5 |  | 1 (12.5%) | 7 |  |
| *France* | 4 (12.9%) | 6.75 |  | 1 (3.23%) | 30 |  |
| *Germany* | 7 (29.17%) | 2.43 |  | 1 (3.85%) | 25 |  |
| *Ghana* | 0 (0%) | (Note 1) |  | 0 (0%) | (Note 2) |  |
| *Greece* | 0 (0%) | (Note 1) |  | 0 (0%) | (Note 2) |  |
| *India* | 0 (0%) | (Note 1) |  | 0 (0%) | (Note 2) |  |
| *Iran, Islamic Rep.* | 0 (0%) | (Note 1) |  | 0 (0%) | (Note 2) |  |
| *Ireland* | 0 (0%) | (Note 1) |  | 0 (0%) | (Note 2) |  |
| *Israel* | 0 (0%) | (Note 1) |  | 1 (12.5%) | 7 |  |
| *Italy* | 6 (14.29%) | 6 |  | 3 (8.33%) | 11 |  |
| *Japan* | 5 (10%) | 9 |  | 3 (6%) | 15.67 |  |
| *Korea, Rep.* | 1 (14.29%) | 6 |  | 1 (16.67%) | 5 |  |
| *Malaysia* | 0 (0%) | (Note 1) |  | 0 (0%) | (Note 2) |  |
| *Netherlands* | 10 (24.39%) | 3.1 |  | 6 (14.63%) | 5.83 |  |
| *New Zealand* | 5 (62.5%) | 0.6 |  | 0 (0%) | (Note 2) |  |
| *Norway* | 4 (66.67%) | 0.5 |  | 2 (25%) | 3 |  |
| *Pakistan* | 1 (33.33%) | 2 |  | 1 (33.33%) | 2 |  |
| *Peru* | 0 (0%) | (Note 1) |  | 0 (0%) | (Note 2) |  |
| *Poland* | 1 (33.33%) | 2 |  | 1 (50%) | 1 |  |
| *Portugal* | 1 (100%) | 0 |  | 0 (0%) | (Note 2) |  |
| *Romania* | 0 (0%) | (Note 1) |  | #DIV/0! | (Note 2) |  |
| *Singapore* | 0 (0%) | (Note 1) |  | 1 (50%) | 1 |  |
| *Spain* | 10 (35.71%) | 1.8 |  | 2 (6.67%) | 14 |  |
| *Sweden* | 6 (50%) | 1 |  | 0 (0%) | (Note 2) |  |
| *Switzerland* | 2 (16.67%) | 5 |  | 2 (14.29%) | 6 |  |
| *Thailand* | 1 (50%) | 1 |  | 1 (50%) | 1 |  |
| *United Arab Emirates* | 0 (0%) | (Note 1) |  | 0 (0%) | (Note 2) |  |
| *United Kingdom* | 21 (18.26%) | 4.48 |  | 15 (11.72%) | 7.53 |  |

|  |  |  |  |  |
| --- | --- | --- | --- | --- |
| *United States* | 51 (16.61%) | 5.02 | 45 (15.05%) | 5.64 |

*Note 1:* There were no female authors from these countries. For reference, the number of male authors was as follows: Argentina=2, Bangladesh=2, Colombia=1, Ghana=1, Greece=0, India=14, Iran, Islamic Rep.=1, Ireland=6, Israel=9, Malaysia=2, Peru=0, Romania=1, Singapore=2, and United Arab Emirates=1.
*Note 2:* There were no female authors from these countries. For reference, the number of male authors was as follows: Argentina=2, Bangladesh=2, Colombia=1, Ghana=1, Greece=1, India=15, Iran, Islamic Rep.=1, Ireland=5, Malaysia=1, New Zealand=8, Peru=1, Portugal=1, Romania=0, Sweden=11, and United Arab Emirates=1.

### Binary logistic regression

The year (β=0.069, *p*<0.001), the first author being Hispanic (β=-3.353, *p*=0.003), and the senior author being a female (β=1.124, *p*<0.001) had a significant positive effect, while the senior author being Hispanic has a negative effect on female first authorship (β=-2.466, *p*=0.044) (**Table 4**). The senior author having PhD qualification (β=0.753, *p*=0.021) had a significant positive effect on female first authorship (**Table 4**).

**Table 4.** Results from the logistic regression model

| Predictor | | $\beta$ | S.E. | Wald | df | Sig. | OR | 95% C.I. | |
| --- | --- | --- | --- | --- | --- | --- | --- | --- | --- |
|  |  |  |  |  |  |  |  | Lower | Upper |
| **Year** |  | 0.069 | 0.014 | 24.625 | 1 | **0.000** | **1.071** | **1.043** | **1.101** |
| **Race of First Author** | *White* | *(Ref)* | *(Ref)* | 12.439 | 3 | **0.006** | *(Ref)* | *(Ref)* | *(Ref)* |
|  | *Asian* | -1.103 | 0.567 | 3.783 | 1 | 0.052 | 0.332 | 0.109 | 1.009 |
|  | *Hispanic* | 3.353 | 1.142 | 8.628 | 1 | **0.003** | **28.599** | **3.052** | **267.972** |
|  | *Black* | 22.409 | 40192.969 | 0 | 1 | 1 | 5396767795 | 0 | . |
| **First Author's Education** | *MD* | *(Ref)* | *(Ref)* | 4.508 | 6 | 0.608 | *(Ref)* | *(Ref)* | *(Ref)* |
|  | *Master's* | 1.62 | 1.675 | 0.935 | 1 | 0.334 | 5.052 | 0.19 | 134.625 |
|  | *Master's, PhD* | 21.723 | 40192.969 | 0 | 1 | 1 | 2717326093 | 0 | . |
|  | *MD, Master's* | 0.311 | 0.381 | 0.666 | 1 | 0.414 | 1.365 | 0.646 | 2.884 |
|  | *MD, Master's, PhD* | -19.139 | 28418.283 | 0 | 1 | 0.999 | 0 | 0 | . |
|  | *MD, PhD* | 0.141 | 0.283 | 0.248 | 1 | 0.618 | 1.151 | 0.661 | 2.004 |
|  | *PhD* | 0.631 | 0.35 | 3.258 | 1 | 0.071 | 1.88 | 0.947 | 3.732 |
| **Gender of Senior Author(1)** | *Female* | 1.124 | 0.22 | 26.138 | 1 | **0.000** | **3.078** | **2** | **4.736** |
| **Race of Senior Author** | *White* | *(Ref)* | *(Ref)* | 4.199 | 3 | 0.241 | *(Ref)* | *(Ref)* | *(Ref)* |
|  | *Asian* | 0.207 | 0.551 | 0.141 | 1 | 0.707 | 1.23 | 0.418 | 3.625 |
|  | *Hispanic* | -2.466 | 1.225 | 4.054 | 1 | **0.044** | **0.085** | **0.008** | **0.937** |
|  | *Black* | -42.27 | 56841.443 | 0 | 1 | 0.999 | 0 | 0 | . |
| **Senior Author's Education** | *MD* | *(Ref)* | *(Ref)* | 7.168 | 4 | 0.127 | *(Ref)* | *(Ref)* | *(Ref)* |
|  | *Master's, PhD* | -19.643 | 27906.758 | 0 | 1 | 0.999 | 0 | 0 |  |
|  | *MD, Master's* | 0.619 | 0.46 | 1.812 | 1 | 0.178 | 1.857 | 0.754 | 4.571 |
|  | *MD, PhD* | -0.071 | 0.281 | 0.064 | 1 | 0.8 | 0.931 | 0.537 | 1.614 |
|  | *PhD* | 0.753 | 0.326 | 5.337 | 1 | **0.021** | **2.123** | **1.121** | **4.02** |
| **Constant** | - | -140.293 | 27.907 | 25.273 | 1 | **0.000** | 0 | - | - |

The model explained 12.2% (Cox and Snell R squared) and 18.2% (Nagelkerke R squared) variability in the first authorship.

## DISCUSSION

Over the 22-year study period, we observed a general trend towards an increased representation of female authors in first and senior authorship of gastroenterology and hepatology RCTs published in high-impact journals. However, progress remains slow, and disparities persist. Moreover, racial disparities in the subsample of female authors were similar to the entire study population, indicating the need to address the gender and racial disparities in authorship separately. For instance, the significantly high male-to-female ratio among Asian authors in the first authorship position may warrant targeted gender-related interventions. At the same time, there is an urgent need to increase the representation of Hispanic and Black racial/ethnic groups across both authorship positions.

Although we did not observe statistically significant differences in gender distribution by World Bank income and regional groups, there were statistically significant differences at the country level in the first authorship position. High research output countries in life sciences in 2022 [31], like the United States, United Kingdom, China, and Japan, collectively contributed over half of the female authors in the first authorship position but with a male-to-female ratio ranging from 9 in Japan to 4.48 in the United Kingdom which was well above the median male to female ratio of 3.23 in our study population. Other high-research output countries like Canada, Germany, Netherlands, and Spain had better female representation with a male-to-female ratio below the median.

An important finding of our study, at least in terms of future direction to improve gender representation in gastroenterology and hepatology research output, is the positive association between gender and the education of first and senior authors. Consistent with our observations, Polanco *et al.* [5] and Leung *et al.* [6] noted a higher proportion of females in first authorship positions in hepatology journals when the senior author was female. However, the proportion of female senior authors in gastroenterology and hepatology journals has remained low for at least three decades [8]. Similarly, in an analysis of 73 US gastroenterology fellowship programs, Sethi *et al.* [12] demonstrated that female leadership as the program director and gastroenterology division chief was associated with increased female fellows and female program directors, respectively. Furthermore, a recent survey of gastroenterology fellows by David *et al.* [32] and Advani *et al.* [16] showed that female fellows perceived the absence of same-sex mentors as gender bias in the workplace which was a significant deterrent for them to pursue a career in advanced endoscopy or gastroenterology in general. In another survey of foundation doctors, gender disparity in gastroenterology itself was cited as a deterrent to selecting gastroenterology as a speciality [33].

Nonetheless, the current study confirms the findings of the previous studies on the underrepresentation of senior female authors in gastroenterology and hepatology research publications [5, 6, 8, 9]. The low representation of women in senior academic positions has been attributed to the “leaky pipeline” phenomenon, where fewer women physicians move up the academic hierarchy [34, 35, 36, 37]. However, as argued by Oxentenko *et al.* [38], increasing female representation in the academic pipeline alone may not be effective with the gender parity achieved in medical school enrolments in high research output countries for over a decade [39, 40, 41]. Instead, organisational allyship of female researchers with male allies, mentors and sponsors, sensitisation of male allies to promote women researchers, and support for female researchers to overcome imposter syndrome and microaggressions may be necessary for a systemic change [15, 33, 38].

It is also essential to consider that as many as 40% of gastroenterologists do not desire leadership roles [18]. Among those who do, similar proportions of men and women have commonly perceived barriers such as increased workload, decreased time for personal life, and interference with clinical duties [15, 18] addressing which may require independent system-level interventions and rethinking of work culture such as the introduction of family-friendly policies as exemplified by American Board of Medical Specialties’ minimum six weeks of parental, caregiver and medical leave during residency [42, 43] and part-time employment practices for female gastroenterologists which, in the past, was not favoured due to issues related to high patient load, financial demands of office overhead, and malpractice insurance [15, 19]. Therefore, achieving gender parity in gastroenterology and hepatology research will require a long-term, multifaceted approach involving individuals, academic institutions, professional societies, and national health agencies [44, 45].

### Limitations

The categorisation of race/ethnicity is based on publicly available information and self-identification, which may not capture the full complexity of an individual’s racial or ethnic background. Moreover, our analysis was restricted to RCTs published in leading gastroenterology and hepatology journals which may not be generalisable to other types of publications or research fields.

## CONCLUSION

Although the proportion of women in medicine and academia has increased in the past two decades, our findings highlight the persistent gender and racial differences in the first and senior authorship of gastroenterology and hepatology RCTs from 2000 to 2022 in high-impact journals. Therefore, efforts to promote diversity and inclusion, such as organisational allyship, targeted mentorship programs for female and non-White researchers, and family-friendly policies, should be prioritised to address these disparities and ensure equitable representation in leadership positions and high-impact publications.

## Data Availability

All data produced in the present study are available upon reasonable request to the authors

## Declarations

*Conflicts of interest:* The authors declare no conflicts of interest.

*Ethics statement:* Since this bibliometric study involves publicly available data, approval from the institutional review board was not sought.

*Source of Funding:* This manuscript did not receive any research funding.

*Authors’ contribution statement:* All authors had access to the data and contributed significantly to writing the manuscript.

## Notes

### Competing Interest Statement

The authors have declared no competing interest.

### Funding Statement

This study did not receive any funding

